# Whole-exome sequencing of the mummified remains of Cangrande della Scala (1291–1329 CE) indicates the first known case of late-onset Pompe disease

**DOI:** 10.1101/2021.06.08.21258201

**Authors:** Barbara Iadarola, Denise Lavezzari, Alessandra Modi, Chiara Degli Esposti, Cristina Beltrami, Marzia Rossato, Valentina Zaro, Ettore Napione, Leonardo Latella, Martina Lari, David Caramelli, Alessandro Salviati, Massimo Delledonne

**Author notes:** B.I. and D.L contributed equally.

## Abstract

Mummified remains of relevant historical figures are nowadays an important source of information to retrace data concerning their private life and health, especially when historical archives are not available. Next-generation-sequencing was proved to be a valuable tool to unravel the characteristics of these individuals through their genetic heritage. Using the strictest criteria currently available for the validation of ancient DNA sequences, whole-genome and whole-exome sequencing were generated from the mummy remains of an Italian nobleman died almost 700 years ago, Cangrande della Scala. While its genome sequencing could not yield sufficient coverage for in depth investigation, exome sequencing could overcome the limitations of this approach to achieve significantly high coverage on coding regions, thus allowing to perform the first extensive exome analysis of a mummy genome. Similar to a standard “clinical exome analysis” conducted on modern DNA, an in-depth variant annotation, high-quality filtering and interpretation was performed, leading to the identification of a genotype associated with late-onset Pompe disease (glycogen storage disease type II). This genetic diagnosis was concordant with the limited clinical history available for Cangrande della Scala, who likely represents the earliest known case of this autosomal recessive metabolic disorder.

## INTRODUCTION

DNA is naturally broken down into fragments after death and is ultimately degraded to single nucleotides, but sequence information can be recovered from samples that are hundreds of thousands of years old[1]. If well-preserved biological samples are available, next-generation sequencing technologies can provide information about historical figures from the recent past, helping to clarify aspects of their official and private lives that cannot be resolved using traditional historical sources. DNA analysis is a non-traditional source of historical information, but it facilities objective and accurate historical reconstructions and is therefore an important resource that can be used to support evidence from traditional sources such as documents, literature and artwork.

An interesting case study is Cangrande della Scala (1291–1329 CE), lord of Verona from 1311 to 1329, a great military commander and politician who brought neighboring cities under his control to form a “kingdom” of the Venetian hinterland spanning from Verona to Treviso. Verifiable data about his private life and health are scarce because the Scaliger family archives were destroyed, forcing historians to rely on less reliable sources that may be influenced by positive or negative bias.

Cangrande della Scala was interred in a marble tomb that promoted mummification. His remains were exhumed in 2004 for scientific analysis by a multidisciplinary team of researchers, revealing the presence of digitalis in his well-preserved organs[2],[3]. This led to several hypotheses, including murder by poisoning[4] and the therapeutic use of digitalis to remedy a cardiac disorder[4],[5],[6]. Here we used samples of bone tissue from the mummified remains for the extraction of ancient DNA, followed by clinical whole-genome sequencing (WGS) and whole-exome sequencing (WES). We identified two pathogenic variants in the *GAA* gene encoding *α*-glucosidase, a genotype associated with late-onset Pompe disease (also known as acid maltase deficiency, acid *α*-glucosidase deficiency, and glycogen storage disease type II). The clinical phenotype of this disease is consistent with data from the historical records, suggesting that Cangrande della Scala is the earliest known case of this prototypic lysosomal storage disorder.

## MATERIALS AND METHODS

### Conservation of the biological remains

In 2007, the biological remains of Cangrande della Scala that had not been reinterred were deposited in the Natural History Museum of Verona to be preserved and made available for further analysis. The remains (parts of the liver, phalanges, metatarsal and cuneiform bones) were placed in sterile receptacles and stored in the dark at 19–21 °C and 40–45% relative humidity, with periodic monitoring. These are standard conditions for the storage of biological material[7]. The selection and collection of samples was carried out in the Zoology Laboratory of the Natural History Museum of Verona.

### Sample preparation and DNA extraction

DNA isolation and library preparation were carried out at the Molecular Anthropology and Paleogenetic Laboratory of the Department of Biology, University of Florence, using facilities exclusively dedicated to ancient DNA analysis, following stringent protocols to prevent contamination with present-day DNA[8]. Negative controls were included in each experimental step. The right intermediate cuneiform bone and a small portion of mummified liver tissue were collected for DNA analysis. To remove potential contaminants, the outer layer of the bone sample was brushed with disposable tools and irradiated with ultraviolet light (254 nm) for 45 min in a Biolink DNA Crosslinker (Biometra). Bone powder was then collected from the densest part of the bone using a low-speed dental micromotor equipped with disposable tungsten carbide ball burrs. DNA was extracted from 50 mg of bone powder using a silica-based protocol that allows DNA molecules to be recovered efficiently even if highly fragmented[9]. In the final step, DNA was eluted twice in 50 µl TET buffer (10 nM Tris, 1 mM EDTA, 0.05% Tween-20). DNA was extracted from 50 mg of mummified liver tissue using the QIAamp DNA mini kit according to the manufacturer’s recommendations (Qiagen).

### Whole-genome library preparation and sequencing

Sequencing libraries suitable for Illumina platforms were prepared from 20 µl of DNA extracted from bone or liver tissue following a protocol optimized for ancient samples[10]. The resulting data were used to evaluate the deamination rate to confirm the authenticity of the genetic material. A partial uracil-DNA glycosylase treatment[11] was applied to an additional 30-µl aliquot of DNA extracted from bone. This treatment removes internal uracil residues and abasic sites, reducing the probability of errors during variant calling. A unique combination of two indices per library was used for barcoding. Libraries were sequenced in 150-bp paired-end mode on a NovaSeq 6000 instrument (Illumina) to generate an average 1× coverage of the entire genome.

### Bioinformatics analysis of WGS data: preservation and contamination estimates, molecular sex determination, and mitochondrial genome reconstruction

Sequences were demultiplexed and sorted according to the indices, and raw sequence data from all three libraries were analyzed using an established pipeline[12]. Adapters were clipped-off and reads with a minimum overlap of 10 bp were merged in a single sequence using Clip&Merge v1.7.4. Merged reads were then mapped onto GRCh38 using BWA v0.7.17-r1188[13], setting parameters to improve the accuracy of ancient DNA reads (-l = 16500, -o = 2 and -n = 0.01)[14]. Only reads with a map and base qualities score ≥ 30 were retained. Reads mapped onto the human genome were authenticated by deamination and fragmentation pattern analysis using mapDamage2.0[15].

Molecular sex determination was applied to the bone sample by comparing the number of alignments to the Y chromosome and the total number of alignments to the X and Y chromosomes in the libraries prepared with and without uracil-DNA glycosylase[16]. The mitochondrial genome was reconstructed from the same libraries to assess overall DNA preservation and contamination. Reads mapping to the mitochondrial genome were extracted from BAM files using SAMtools v1.7[17]. For the library prepared without uracil-DNA glycosylase, the Schmutzi pipeline[18] was used to call the consensus sequence and to evaluate the level of contamination with present-day human DNA[19]. For the library prepared using a partial uracil-DNA glycosylase treatment, the consensus sequence was called using mpileup and vcfutils.pl in the SAMtools package. To estimate the ratio of contaminant/authentic DNA in the mitochondrial sequence data, a likelihood-based method was used as previously described[20]. The mitochondrial haplogroups were assigned according to PhyloTree build 17[21] using Haplogrep2[22].

### Exome enrichment and WES

The DNA library prepared from bone was captured using the Twist Bioscience Human Core Exome Kit + RefSeq v1.3 protocol. Single-plex exome capture was carried out using 120-bp biotinylated probes, with minor modifications to the standard protocol due to the high level of sample degradation. All the available material from the library was used, although the DNA input requirements of the standard protocol were higher than the sample’s initial input (300 ng instead of 500 ng). After the washing steps to remove nonspecific targets, the remaining material was eluted in 22.5 µl of water, without keeping the backup slurry. Ten cycles of amplification were performed rather than the eight cycles recommended by the protocol. The final PCR cleanup was carried out using a 1.5× ratio of Twist Bioscience Beads. The enriched library was validated using a Tape Station 4150 High Sensitivity D1000 assay kit (Agilent Technologies) and quantified by RT-PCR using the Lib Quant kit (Roche). WES was then performed on a NovaSeq 6000 instrument in 2× 100-bp paired-end mode.

### Bioinformatics analysis on WES data: read alignment, variant calling, and data authentication

The WES FASTQ files were quality checked using FastQC (http://www.bioinformatics.babraham.ac.uk/projects/fastqc/). Adapters and low-quality bases were removed and reads were aligned with the human reference genome (GRCh38/hg38) using the Paleomix bam_pipeline v1.2.13.8[23] with BWA-mem v0.7.17[13] and a disabled “--collapse” parameter to properly calculate the insert size for all the sequenced fragments. Duplicated reads were removed using Picard MarkDuplicates v2.21.1. GATK Base Recalibrator v4.1.8.1[24] and BamUtil clipoverlaps v1.4.14 were then applied to adjust base quality and soft-clip overlapping reads. Coverage and genotypability metrics were calculated using CallableLoci in GATK v3.8. The FOLD80 penalty value was calculated using Picard CollectHSMetrics v2.21.1 (http://broadinstitute.github.io/picard/). Variants were identified using GATK HaplotypeCaller v4.1.8.1 (with parameter “--dont-use-soft-clipped-bases” set to “true”), producing a gVCF file. Variants were then recalibrated and filtered by following the GATK Hard Filtering Best Practices.

The authenticity of the WES data was estimated as previously described for WGS. Additionally, nuclear contamination was estimated by measuring the heterozygosity of the X chromosome[25] using the ANGSD pipeline[26]. Because males have only one copy of the X chromosome, any heterozygosity on this chromosome in males indicates contamination.

### Variant annotation, prioritization and phasing

The gVCF file was annotated using Golden Helix VarSeq v.2.2.1 (Golden Helix) and the following databases: ClinVar and HGMD Professional v2020.1[27] were used to investigate the clinical significance of identified variants, whereas the population frequency databases of 1000Genomes Project Phase3, gnomAD v2.0.1 and ESP6500 v2 were used to determine the frequency of variants. Similarly, an internal database was used to flag rare Italian variants. Several prediction tools were used to calculate the pathogenicity scores for each genetic variation (FATHMM, GERP, Polyphen, SIFT, PhastCons and PhyloP) and the RefSeq Genes database was integrated to provide the effect of each variant. Variants were then prioritized using two pipelines. The “annotated variants” pipeline retained only those variants classified as “Pathogenic”, “Likely Pathogenic”, “Conflicting”, “Uncertain Significance” or “Other” in ClinVar, or classified as “DM” or “DM?” in HGMD. Variants with an alternative allele frequency below 5% in the population frequency databases were flagged. The “predicted coding variants” pipeline retained variants present in exonic or splicing site regions but without reported clinical significance in ClinVar and HGMD. Variants with an alternative allele frequency below 1% in the population frequency databases were flagged, focusing on those with: (1) a “LOF”, “Missense” or “Splice_region_variants” effect in the RefSeq database; (2) a predicted “Damaging” effect by three or more prediction tools applied to the dbNSFP database (SIFT, Polyphen2, MutationTaster, MutationAssessor and FATHMM); and (3) a frequency below 2% in the Functional Genomics Variant database. Phasing of WES reads was accomplished using Eagle v2.4.1, with the provided hg38 genetic map and the reference panel from gnomAD v3.1 “HGDP+1KG callset” (https://gnomad.broadinstitute.org/downloads#v3-hgdp-1kg).

## RESULTS

### WGS performance and authentication of ancient DNA

DNA extracted from the mummified remains of Cangrande della Scala (right intermediate cuneiform bone and liver) was used for three exploratory WGS experiments. Two WGS datasets were prepared from bone DNA, one including partial uracil-DNA glycosylase treatment and one with no treatment. A third dataset was prepared from liver DNA to assess the degree of DNA preservation. The percentage of human sequence content in all three samples was compared by alignment to the human reference genome (Table 1). This revealed an extremely low percentage of mapped fragments from the liver sample (0.06%) but a much higher percentage from the bone sample (23%).

**Table 1.** Exploratory WGS analysis, indicating the human sequence content in three DNA samples. *The table shows the number of sequenced reads, the GC percentage, the number of mapped reads before and after removing PCR duplicates, the percentage of fragments mapped to the human reference genome, and the average coverage of the genome.*

| Sample ID | No. of merged reads | GC content (%) | No. of mapped reads before removing PCR duplicates | No. of mapped reads after removing PCR duplicates | Human DNA (%) | Mean coverage (×) |
| --- | --- | --- | --- | --- | --- | --- |
| Cangrande liver | 4,445,677 | 34.55 | 2,572 | 494 | 0.06 | 0.00 |
| Cangrande bone no UDG | 7,924,124 | 41.85 | 1,856,551 | 1,533,864 | 23.43 | 0.0392 |
| Cangrande bone | 6,384,132 | 41.66 | 1,488,301 | 1,099,139 | 23.31 | 0.0289 |

The fraction of human DNA recovered from the cuneiform bone sample was consistent with values obtained from other small foot bones (phalanxes) in human remains of similar age[28]. We carried out several tests to confirm the ancient nature of the sequenced human DNA and to exclude modern DNA contamination. The tests revealed that the library sequences showed features typical of degraded DNA, such as an average length of ∼90 bp without uracil-DNA glycosylase treatment and 86 bp with partial uracil-DNA glycosylase treatment. Furthermore, the frequency of 5′ cytosine deamination in the library prepared without uracil-DNA glycosylase treatment was consistent with the age of the sample[29] (Supplementary Table S1, Supplementary Figures S1 and S2). Complete mitochondrial genomes could be reconstructed from both libraries (mean coverage of 11.42× and 8.22× respectively) and both sequences were unambiguously assigned to the same mitochondrial haplogroup (X2b11) with a maximum score (Supplementary Table S2). Finally, present-day human DNA contamination in mitochondrial sequences, estimated using two different methods, did not exceed 1% (Supplementary Table S2). Genetic sex determination revealed Ry values of 0.0885 and 0.0887 for the libraries prepared with and without uracil-DNA glycosylase treatment, respectively, confirming that the bone DNA belonged to a male individual (Supplementary Table S3).

### Exome capture and WES performance

Having confirmed satisfactory DNA preservation, we used the bone sample for WES to enrich for human sequences and achieve good coverage for variant calling at a reasonable cost. To limit the effect of DNA damage on variant calling, whole-exome capture was restricted to the library prepared using partial uracil-DNA glycosylase treatment. Quality control of the enriched library revealed a size range of 116–769 bp (average = 277 bp) and a concentration of 4.68 ng/µl (Figure 1).

**Figure 1.**
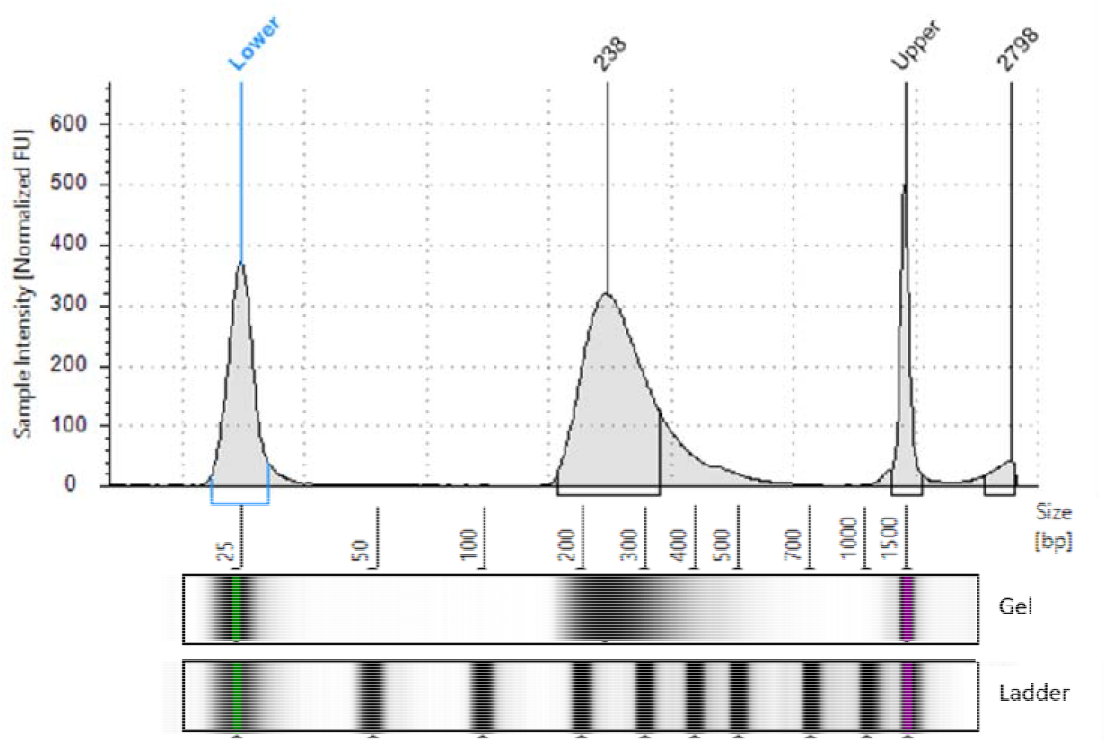
Distribution of fragment sizes in the enriched library used for WES.

The WES library was sequenced twice, yielding 62 million and 77 million fragments, respectively (Table 2). The mapped coverage on the exome target was on average 29.35× in the first run, with a large proportion of PCR duplicates (51.72%). This increased slightly to 31.83× in the second run, but again the low coverage reflected the large proportion of PCR duplicates (55%). When the two runs were combined, the mapped coverage was 37.28× and the percentage of duplicates increased to 65%, indicating that library saturation had been achieved. Despite the low average coverage, the high uniformity of enrichment (FOLD80 penalty ≥ 1.41) allowed us to cover 99.16% of the exome target with at least 10 reads (Twist design) and to genotype 93.38% of the target bases (Table 2).

**Table 2.** Performance of the two WES runs. *The table shows the number of sequenced fragments, the number of mapped reads before and after duplicate removal, the percentage of duplicated reads, the average coverage of the target, the percentage of the target covered at least by 10 reads, the genotypability, and the uniformity of coverage (FOLD80 penalty value).*

|  | No. of fragments | No. of mapped reads before removing PCR duplicates | Percent dupl. | No. of mapped reads after removing PCR duplicates | Mean coverage (×) | %10× | %PASS | FOLD80 |
| --- | --- | --- | --- | --- | --- | --- | --- | --- |
| **RUN1** | 62,394,319 | 107,031,260 | 51.72 | 51,669,825 | 29.35 | 98.08 | 93.66 | 1.53 |
| **RUN2** | 77,921,888 | 135,479,585 | 55.99 | 59,615,288 | 31.83 | 98.60 | 93.60 | 1.39 |
| **MERGED RUNS** | 140,316,207 | 242,510,843 | 65.47 | 83,738,060 | 37.28 | 99.16 | 93.38 | 1.41 |

### Exome data authentication and estimate of nuclear DNA contamination

The misincorporation patterns and fragment sizes of the WES data (Supplementary Figure S3) were consistent with the profiles derived from the WGS data (Supplementary Figure S2): 2.17% C > T transitions at the first 5’ base and an average fragment length of 100 bp, approximately 10 bp longer than the pre-capture size, in agreement with earlier studies[9],[30],[31] (Supplementary Table S4). The WES data were also used to estimate putative nuclear DNA contamination. Taking advantage of the male genetic sex of the sample, we measured the heterozygosity observed at 1344 polymorphic sites on the X-chromosome. This revealed 0.0047% X-chromosome contamination, with an estimated error of 2.177062 × 10^−3^ thus confirming the authenticity of the ancient DNA data (Supplementary Table S4).

### Variant identification and prioritization

We identified 24,769 variants in the target regions, 34.15% of which were homozygous (33% SNVs and 1% INDELs) and 65.85% of which were heterozygous (63% SNVs and 2% INDELs). The overall Ti/Tv ratio was 2.77, which is consistent with other WES studies[32]. The variants were then investigated to determine whether Cangrande della Scala carried mutations that may have contributed to his death. Variants were prioritized using two dedicated pipelines (annotated variants and predicted coding variants, as described in the methods section). Among the final set of prioritized variants (249 annotated variants and 1342 predicted coding variants, Supplementary Tables S5 and S6), those classified in HGMD and/or ClinVar were investigated in detail. This reduced the priority list to 210 ClinVar variants and 179 HGMD variants, 140 of which were common to both databases. Subsequent analysis focused on 113 rare clinical variants associated with more severe diseases. They included two heterozygous missense mutations in two exons of the *GAA* gene associated with clinical symptoms consistent with the ones reported for Cangrande’s death. The first variant (c.1465G>A) was classified as “pathogenic/likely pathogenic” in ClinVar and damaging (DM) in HGMD, whereas the second (c.271G>A) was classified as “benign/likely benign” in ClinVar and likely damaging (DM?) in HGMD. Both variants are associated with the autosomal recessive phenotype “late-onset Pompe disease”. None of the other variants were associated with phenotypes of interest consistent with Cangrande’s chronic illness and death. Nevertheless, given the conflicting interpretations of the second variant, we investigated other mutated genes correlating with the regulatory activity of lysosomal enzymes to identify any combined effects of multiple genetic causes for the same phenotype. Another six rare exon missense variants were present in the *ATP6* gene, four of which were defined as damaging in the database of human nonsynonymous SNPs and their functional predictions (dbNSFP). From the set of common variants, two synonymous exon variants were detected in the *TFEB* gene.

### Phasing the GAA variants

Chromosome-wide phasing was used to determine the cis/trans phase of the two heterozygous genotypes in the *GAA* gene, considering all the variants on chromosome 17. Only one of the two variants of interest (c.271G>A) could be correctly phased, whereas the other (c.1465G>A) was discarded by the algorithm because it was missing from the reference population. To exclude the possibility that the two variants did not constitute a haplotype, we assessed the longest haplotype in the reference population carrying variant c.271G>A. The aim was to confirm that the haplotype containing c.271G>A spanned over the c.1465G>A variant position, which could be inferred as a reference for this haplotype in the population. This allowed the identification of a set of variants consistently inherited together with c.271G>A in 98% of reference individuals, namely in a linkage disequilibrium block more than 14 kb in length (chr17:80,096,549–80,110,889) and spanning over the c.1465G>A variant position. It is therefore very unlikely that the latter variant was inherited with c.271G>A on the same chromosome. Moreover, the frequency of the two variants differed in the general population frequency databases, confirming the low probability of both being inherited together (c.271G>A allele frequency = 3% in all the population databases, c.1465G>A allele frequency reported in two databases only, with values of 0.0009% and 0.0015%, respectively). These data support the hypothesis of a compound heterozygous genotype (variants affect the *GAA* gene in trans) and increase the likelihood of a diagnosis of late-onset Pompe disease.

## DISCUSSION AND CONCLUSION

We have described the successful clinical analysis of a 700-year-old human mummy by WES and demonstrated that exome enrichment applied to ancient human DNA can lead to a genetic diagnosis that may help to support historical data. By integrating WGS and WES, we confidently assessed the authenticity of the data and excluded significant bias caused by contamination with modern human DNA.

Next-generation sequencing for the study of ancient samples has mainly targeted small regions, such as the mitochondrial DNA (mtDNA), the Y-chromosome DNA (Y-DNA)[33] or specific SNPs of interest[34],[35]. Only a few “high-quality” ancient human WGS studies have been reported, including a Denisovan[36] (30× mean coverage) and two Neanderthal individuals (52× and 27× mean coverage)[37],[38] investigated mainly to discover gene flow events and admixtures of archaic hominins. Typically, low-coverage genomes are used to investigate genetic diversity at the population level and provide phylogenetic information[39],[40]. Considering the low quantity of human material present in Cangrande’s samples, which would have required an abnormal sequencing effort, we considered WES as a much more affordable technology to eliminate environmental DNA contaminants and enrich for protein-coding regions, which are mainly responsible for the development of Mendelian disorders[41]. WES has already been applied in a limited number of studies, although these did not focus on the functional consequences of genetic variants and their association with clinical phenotypes[42],[43]. WES performed on samples from the mummified remains produced 37× coverage of the protein-coding regions. The genotypability values and number of identified variants were almost comparable to the WES analysis of modern humans[44]. We identified two clinically relevant compound heterozygous variants in the *GAA* gene associated with late-onset Pompe disease, which combined with the effect of two synonymous variants in the *TFEB* gene and six rare missense variants in the *ATP6* gene could very well explain Cangrande’s chronic disease and death.

Contemporaneous sources on Cangrande’s life provide little verifiable information about his health. These documents are also difficult to interpret because the authors were either very close to the court or enemies of the Scaligeri. Most accounts of Cangrande’s childhood are laudatory and are based on literary *topoi*. They describe a child who did not like traditional games or the company of peers, but was predisposed to military life. We know from the pro-Scaliger writer Ferreto Ferreti and from the Paduan enemy Albertino Mussato that Cangrande, even in combat on horseback, preferred to use the bow in the Parthian manner rather than the spear or sword, allowing his arm more freedom of movement[45],[46]. Cangrande was reportedly ill for some time at the age of 23 but recovered enough for battle after imbibing a small dose of an antidote (not better defined) and a sip of wine[47]. The enemy chronicler Mussato attributed the discomfort to one foot, which prevented the Scaligero from riding[46]. He was forced to abandon his horse and accept the draft horse of a peasant[47]. At the age of 29, Cangrande was reportedly pierced by an arrow in the thigh[48], but managed to return to camp, rally his troops and return to fight[49]. The autopsy of Cangrande in 2004 did not reveal any wounded limbs, suggesting that Cangrande instead suffered crippling thigh discomfort, possibly a cramp. In his next battle he once again had to abandon his horse and accept one from a peasant[50]. At the age of 34, Cangrande fell seriously ill for a long time and was given up for dead[51]. Cangrande died at the age of 38, after showing symptoms of malaise for 3 days identified as fever and a generic fluxum, which can be translated as vomiting or perhaps hemorrhage[50]. Some sources report fluxus ventris or intestinal disease with diarrhea, which was disproven in the 2004 autopsy by the discovery and examination of solid fecal matter at the base of the rectum. Cangrande retained his mental clarity at the end of his life, enabling the completion of an important juridical document[51].

The clinical spectrum (infantile, juvenile and adult-onset Pompe disease) is a continuum, depending on the residual activity of *α*-glucosidase. In late-onset forms, the storage of glycogen is confined to the skeletal muscle, heart and liver. The clinical manifestation includes skeletal muscle weakness, respiratory distress due to diaphragm and accessory respiratory muscle weakness, muscle cramps, spontaneous bone fractures and cardiopathy (but normal intelligence). This is entirely consistent with the Cangrande’s three episodes of severe weakness after exercise as reported in the historical records, and his death after 3 days of sickness, but with no mental impairment. No scar was found on his thighs, supporting the hypothesis that the arrow wound reported in 1320 was in fact a severe muscle cramp. Given the finding of digitalis in his body[4], it is possible that it was administered to counteract tachycardia, a key symptom of cardio-respiratory insufficiency, and would represent the first clinical use of this drug.

The first variant (c.1465G>A) found in Cangrande’s *GAA* gene is described in patients with late-onset Pompe disease and inhibits *α*-glucosidase maturation and activity[52]. The second variant (c.271G>A) showed conflicting clinical significance. This allele, known as *GAA*\*2, is frequent in the Caucasian English population (3%). Its biochemical phenotype shows reduced activity toward the natural substrate (glycogen) but normal activity toward the artificial substrate 4-methylumbelliferil-*α*-glucopyranoside. The K_m_ of *α*-glucosidase for glycogen is 10-fold higher in the *GAA*\*2 variant compared to the wild-type enzyme[53]. To explain the absence of this frequent allele in their 15 late-onset cases, the authors cite a modification of the Michaelis-Menten equation[54] demonstrating that the increase in K_m_ is offset by an increase in substrate concentration, restoring the normal turnover rate and establishing an equilibrium substrate concentration below the solubility of glycogen, the critical threshold at which uncontrolled glycogen accumulation might occur. The authors therefore did not class variant *GAA*\*2 as a disease-causing allele, based on the assumption that lysosomal pathology is due to encumbrance. However, lysosomes are involved in many fundamental processes, such as secretion, plasma membrane repair, signaling, energy metabolism, and apoptosis. Two genes have been shown to mediate this feedback mechanism: *TFEB* (enhancing the production of the enzyme) and *ATP6* (controlling the pH of the lysosomal compartment)[55],[56]. We found only two synonymous exon variants in *TFEB*, but six rare missense variants in *ATP6*. The possibility that a compound effect involving the *GAA*\*2 phenotype and another gene or an environmental factor might lead to disease cannot be excluded[53]. We therefore conclude that the additive effects of c.1456G>A and *GAA*\*2 in the *GAA* gene together with *ATP6* variants led to a case of late-onset Pompe disease in Cangrande della Scala.

## Data Availability

The WES variants data are available for download from our public repository using the provided link (VCF file with associated BED files of callable regions).

http://ddlab.sci.univr.it/files/Cangrande/Cangrande.tar.gz

## ACKNOLEDGEMENTS

This research was carried out in the framework of the Joint Project 2018 “The genome of Cangrande della Scala: DNA as historical source”.

## AUTHOR CONTRIBUTIONS

Conceptualization, M.D., L.L., E.N.; methodology, D.C., M.L. M.D.; resources, L.L.; software, B.I., D.L.; validation, A.S.; formal analysis, B.I., D.L, A.S., A.M, V.Z.; investigation, C.D.E, C.B.; writing—original draft, B.I., D.L, M.D., A.S.; writing—review and editing, E.N., M.L., D.C., A.S., M.R., M.D.; supervision, M.R., M.D., A.S.; funding acquisition, L.L and M.D.

## DATA AVAILABILITY

The WES variants data are available for download from our public repository using the link: http://ddlab.sci.univr.it/files/Cangrande/Cangrande.tar.gz (VCF file with associated BED files of callable regions).

