## Supplementary Figures for "Whole-exome sequencing of the mummified remains of Cangrande della Scala (1291–1329 CE) indicates the first known case of late-onset Pompe disease"

**Supplementary Figure S1. Properties of the library prepared from bone DNA without uracil-DNA glycosylase treatment.**

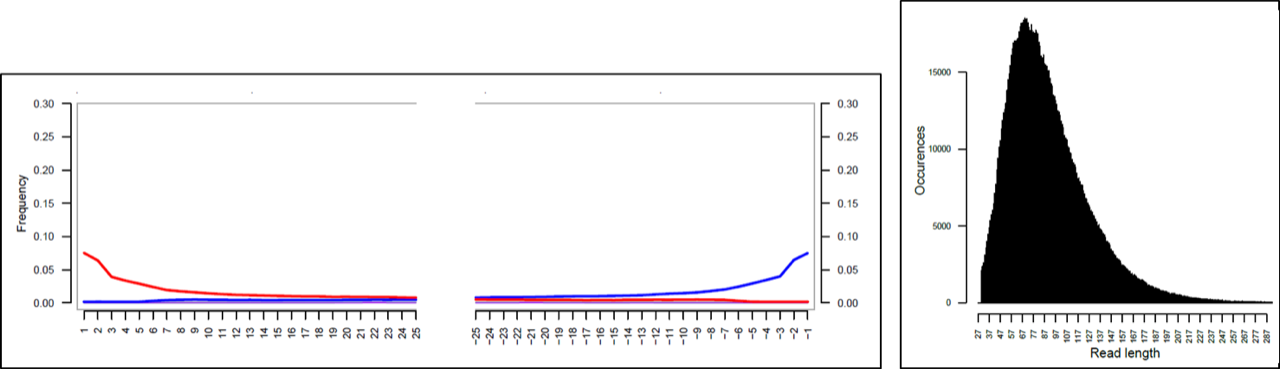

**Supplementary Figure S2. Properties of the library prepared from bone DNA with partial uracil-DNA glycosylase treatment.**

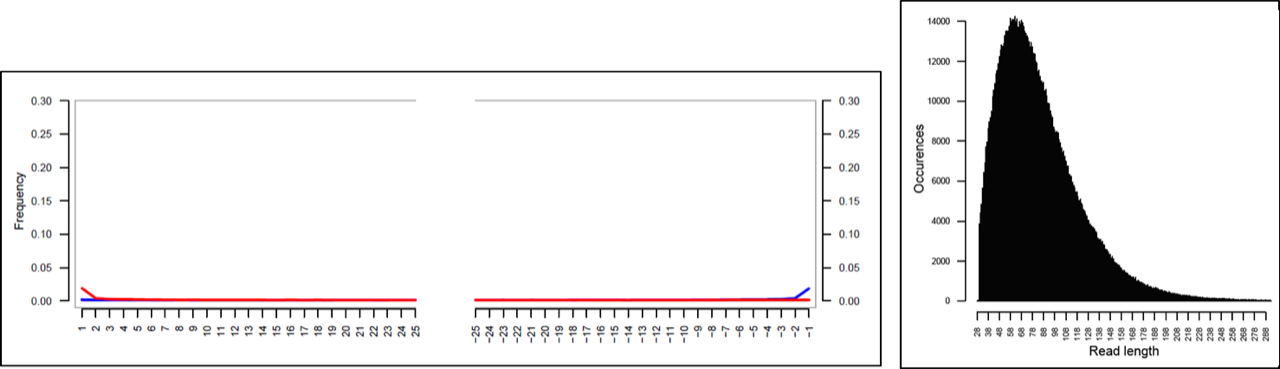

**Supplementary Figure S3. Properties of the exome library.**

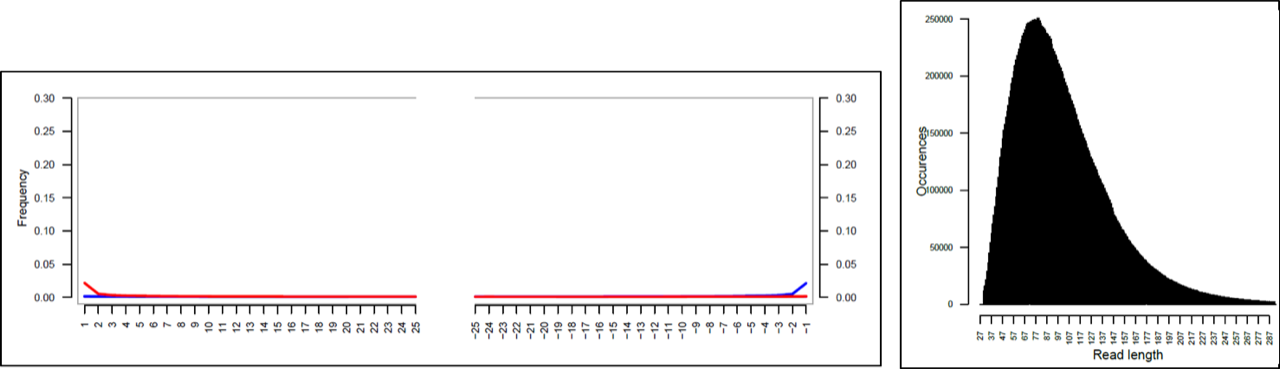
